# Boys are more likely to be undernourished than girls: A systematic review and meta-analysis of sex differences in undernutrition

**DOI:** 10.1101/2020.09.19.20196535

**Authors:** Susan Thurstans, Charles Opondo, Andrew Seal, Jonathan C Wells, Tanya Khara, Carmel Dolan, André Briend, Mark Myatt, Michel Garenne, Rebecca Sear, Marko Kerac

## Abstract

**Background:** Excess male morbidity and mortality is well recognised in neonatal medicine and infant health. In contrast, within global nutrition, it is commonly assumed that girls are more at-risk of experiencing undernutrition. We aimed to explore evidence for any male/female differences in child undernutrition using anthropometric case definitions and the reasons for differences observed.

**Methods:** We searched: Medline, Embase, Global health, Popline and Cochrane databases with no time limits applied. Eligible studies focused on children aged 0-59 months affected by undernutrition where sex was reported. In the meta-analysis, undernutrition-specific estimates were examined separately for wasting, stunting and underweight using a random effects model.

**Results:** 76 studies were identified: 46/76 studies were included in the meta-analysis. In 20 which examined wasting, boys had higher odds of being wasted than girls (pooled OR 1.26, 95% CI 1.13-1.40). 39 examined stunting: boys had higher odds of stunting than girls (pooled OR 1.31 95% CI 1.24-1.39). 25 explored underweight: boys had higher odds of being underweight than girls (pooled OR 1.19, 95% CI 1.07-1.32). There was some limited evidence that the female advantage indicating lower risk of stunting and underweight was weaker in South Asia than other parts of the world.

44/76 (58%) studies discussed possible reasons for boy/girl differences; 11/76 (14%) cited studies with similar findings with no further discussion; 21/76 (28%) had no sex difference discussion. 6/44 studies (14%) postulated biological causes, 21/44 (48%) social causes and 17/44 (38%) to a combination.

**Conclusion:** Our review indicates that undernutrition in children under 5 is more likely to affect boys than girls, though the magnitude of these differences varies and is more pronounced in some contexts than others. Future research should further explore reasons for these differences and implications for nutrition policy and practice.

**Key Questions:** *What is already known?:*

- Undernutrition (wasting, stunting, and underweight) is a public health problem affecting millions of children aged under 5 years globally.
- Although higher neonatal and infant morbidity/mortality for boys is well described, little attention has been given to sex differences in the field of undernutrition due to an assumption that girls are very often disadvantaged over boys.

*What are the new findings?:*

- In most settings studied, undernutrition is more common among boys than girls, though the extent of these differences varies and is reversed in a few contexts.
- Both biological and social mechanisms have been proposed to be responsible for the observed differences as well as a combination of the two.

*What do the new findings imply?:*

- Greater awareness of actual sex differences is needed within the field of nutrition. While sex-specific data is routinely analysed and reported in nutrition surveys it should be used in nutrition programming to better identify and understand what differences exist. Analysis should assess if the sex balance in programme admissions is reflective of the population undernutrition burden.
- Further research is needed to understand the mechanisms that lead to sex and gender differences in undernutrition and their implications. Better epidemiological understanding is a priority, as is work to explore their consequent effects on morbidity and mortality.

## Introduction

Undernutrition is a serious public health problem affecting millions of children worldwide. Recent estimates indicate that stunting (low height-for-age) affects approximately 149 million children, with consequences for growth and cognitive development. Wasting (low weight-for-length), a life-threatening consequence of acute nutrient deficits and/or disease affects over 49 million children globally; 17 million of these are severely wasted. [1] However, these numbers are based on prevalence estimates meaning true numbers may be considerably higher when incidence is taken into consideration. [2]

Sex (referring to biological attributes) and gender (referring to socially constructed roles, behaviours and identities) [3] are important considerations in the public health domain and receive different prominence in different academic and professional fields. Despite considerable research on childhood sex differences in neonatal and infant health, different disciplines often hold surprisingly contrary views on the relative vulnerability of male and female children.

In neonatal medicine and infant health communities, excess male morbidity and mortality is almost universally reported and is widely recognised. [4, 5] Boys are known to be more vulnerable than girls, from as early as the point of conception. [6] Conditions common in childhood such as lower respiratory infections, diarrhoeal diseases, malaria, and preterm birth are all more common in boys than in girls, with the exception of measles, whooping cough and tuberculosis. [7] All of these are not only causes of death but also of weight loss, growth faltering or severe undernutrition among young children. [8] Boy-girl differences have not been explored in detail within the nutrition field, but girls are often widely viewed as more disadvantaged and vulnerable [9] from a gender perspective [10-13].

How underlying biological mechanisms related to sex and social differences in gender translate into the risk of anthropometric deficits and related morbidity and mortality in the field of nutrition remains understudied. Likewise, the practical implications of these differences remain to be determined. In terms of growth and nutrition status, sex differences have long been recognised and reflected through growth charts targeted at individual sexes. [14, 15] On average, boys are slightly heavier and longer at birth and throughout infancy compared to girls, and more detailed studies have shown that the extra average weight of boys is primarily lean mass, whereas fat mass is more similar between the sexes. [16, 17] To evaluate growth and nutritional status therefore, raw anthropometric data are conventionally converted to indices (e.g. weight-for-age; weight-for-length, length-for-age) and expressed in comparison to reference populations as z-scores (standard deviation scores, whereby +1 and −1 z-scores are one standard deviation above and below the reference population median respectively). Data published by WHO in 2006 represent a ‘gold standard’ of how children should grow and were developed from a globally representative reference population of healthy, breastfed children. In constructing the growth standards, data for boys and girls were analysed separately [15] and the resulting growth charts should already therefore account for any sex differences. What has received little attention to date is whether sex differences reappear when z-scores are shifted away from the healthy reference range, which would indicate sex differences in susceptibility to undernutrition.

The objectives of this review were to systematically review the evidence for sex differences in undernutrition in children aged under 5 years, to explore evidence of any male/female differences in child undernutrition, and to document reasons given for any observed differences.

## Methods

This systematic review was conducted following the Preferred Reporting Items for Systematic reviews and Meta-Analyses (PRISMA) guidelines. [18] A protocol for the review was defined, including inclusion and exclusion criteria, and was shared among authors for consensus. The protocol was then registered with the PROSPERO International prospective register of systematic reviews (CRD42018094818).

### Search strategy

Our search strategy captured the concepts of undernutrition, sex and gender. Detailed search terms are in Box 1.

**Box 1. Search terms**

1. undernutrition.mp. (5708)
2. malnutrition.mp. (39279)
3. malnutrition/ or exp fetal nutrition disorders/ or exp refeeding syndrome/ or exp severe acute malnutrition/ or exp kwashiorkor/ or exp starvation/ or exp wasting syndrome/ (25202)
4. (severe adj2 malnutrition).mp. (2131)
5. stunting.mp. (3456)
6. exp Growth Disorders/ (30538)
7. chronic malnutrition.mp. (519) 8. stunt*.mp. (6655)
8. MUAC.mp. (407)
9. mid upper arm circumference.mp. (771)
10. exp Nutritional Status/ (38539)
11. marasmus.mp. or Protein-Energy Malnutrition/ (7366)
12. famine.mp. (1726)
13. exp Starvation/ (9562)
14. (failure adj2 thrive).mp. (5307)
15. 1 or 2 or 3 or 4 or 5 or 6 or 7 or 8 or 9 or 10 or 11 or 12 or 13 or 14 or 15 (123406)
16. limit 16 to (“all infant (birth to 23 months)” or “newborn infant (birth to 1 month)” or “infant (1 to 23 months)” or “preschool child (2 to 5 years)”) (35919)
17. (boy* or girl* or male* or female* or gender or sex).ti,ab. (177252)
18. 17 and 18 (6631)

(Numbers in parenthesis show the number of search results for each line)

Studies were identified by searching the Medline database using the above terms which were then adapted to Embase, Global health, Popline and Cochrane databases. No limits were applied for year of publication in order to capture historical publications on the subject. Studies were restricted to those that included terms for boy, girl, male, female, gender, or sex in the title or abstract, with the aim of filtering through the large body of literature that exists for undernutrition and capturing studies which either directly focused on sex and/or gender in the context of undernutrition or those which disaggregated and reported on it within main findings. As per the PRISMA recommendations, the search strategy was peer reviewed by a librarian.

### Eligibility criteria

Studies were included in the review if they met the following criteria: human studies, 0-59 months of age, male and female participants, and including a focus on undernutrition as a primary outcome and/or undernutrition related mortality and morbidity. Studies of children over 59 months, non-English language, animal studies and studies on overweight/obesity and micronutrient deficiencies were excluded. Both peer reviewed and grey literature were selected. Where studies included data for children both under and over 59 months, where possible, we extracted the data for children <59 months only. Where this was not possible, studies were excluded.

### Data extraction

All records identified through the search were exported into Endnote (Endnote X8, Clarivate analytics). Duplicates were identified and removed. Initial screening was conducted by reading titles and abstracts to identify and remove studies which clearly did not fit our scope. Detailed review of the full text of all remaining results was conducted to determine which met the inclusion and exclusion criteria. When it was not clear how to classify an article, this was resolved through discussion and consensus with two or more authors.

A data extraction template was piloted on a small number of articles before being finalised. Data was extracted on study characteristics and outcomes of interest. These included aims and types of studies, sample size, prevalence and male/female odds ratios for undernutrition, and explanations offered for any differences identified and future recommendations made.

### Analysis

Due to variations in type of paper and study design, the analysis was conducted in two parts: a qualitative systematic review followed by a meta-analysis. Studies were included in the meta-analysis if they focused on stunting, wasting or underweight as an outcome, and presented extractable data that was fully disaggregated by sex.

We performed random effects meta-analyses to pool estimates from studies that included a measurement of undernutrition prevalence, or which assessed risks and determinants of undernutrition, and stratified results by sex. Missing counts, denominators and effect estimates such as odds ratios, relative risk and their associated confidence intervals were calculated from other information provided where it was possible to do so. Studies that presented only adjusted odds ratios or risk ratios were excluded given that studies were likely to adjust for different factors and such adjusted effect estimates were not directly comparable. Undernutrition-specific estimates were pooled separately for wasting, stunting and underweight using a random effects model. Analysis was also stratified by age and country. Pooled effects are presented as odds ratios (OR) and 95% confidence intervals (CI). Meta-regression was conducted to assess the heterogeneity of estimates. Statistical analysis was conducted using Stata version 15.1 (StataCorp. 2017. Stata Statistical Software: Release 15. College Station, TX: StataCorp LLC).

In all studies conducted earlier than 2006, NCHS growth [19] references had been used. In all post 2006 studies that were included, the WHO (2006) growth standards for wasting, stunting and underweight, as measured through standard deviation from the mean z-scores were used. Wasting was defined by weight –for-height z-score <-2; stunting was defined by height-for-age z-score <-2; underweight was defined by weight-for-age z-score <-2

### Risk of bias assessment

We adapted the National Heart, Lung and Blood institute study quality assessment tools for Observational cohort and cross-sectional studies to assess the quality of studies [20], and applied it to studies identified for the meta-analysis. Using this tool, we assessed data sources, a study’s presentation of aims and objectives and target populations, the appropriateness of anthropometric methods and the presentation of results. We adapted the tool to assess if studies acknowledged sex differences in the discussion of results.

### Patient and Public involvement

The design of this review meant it was not appropriate or possible to involve patients or the public in the design, or conduct, or reporting, or dissemination plans of our research.

## Results

### Study selection

The study flow chart in figure 1 summarises our process of identifying studies. The final search of Embase, Global health, Popline and Cochrane databases (conducted in March 2020) identified 34,270 studies, including both peer reviewed studies and grey literature. In addition, 21 studies were found from other sources. After removing duplicates, 22,359 studies remained. Initial screening excluded 21,925 studies as they were unrelated to our review questions. Full texts of the 434 remaining studies were reviewed in detail to assess eligibility. At this stage, a further 358 studies were discarded as they did not meet the inclusion criteria. Mostly because there was no mention of sex or gender in relation to undernutrition. Finally, 76 studies were included in the qualitative synthesis and 46 studies were included in the meta-analysis.

**Figure 1.**
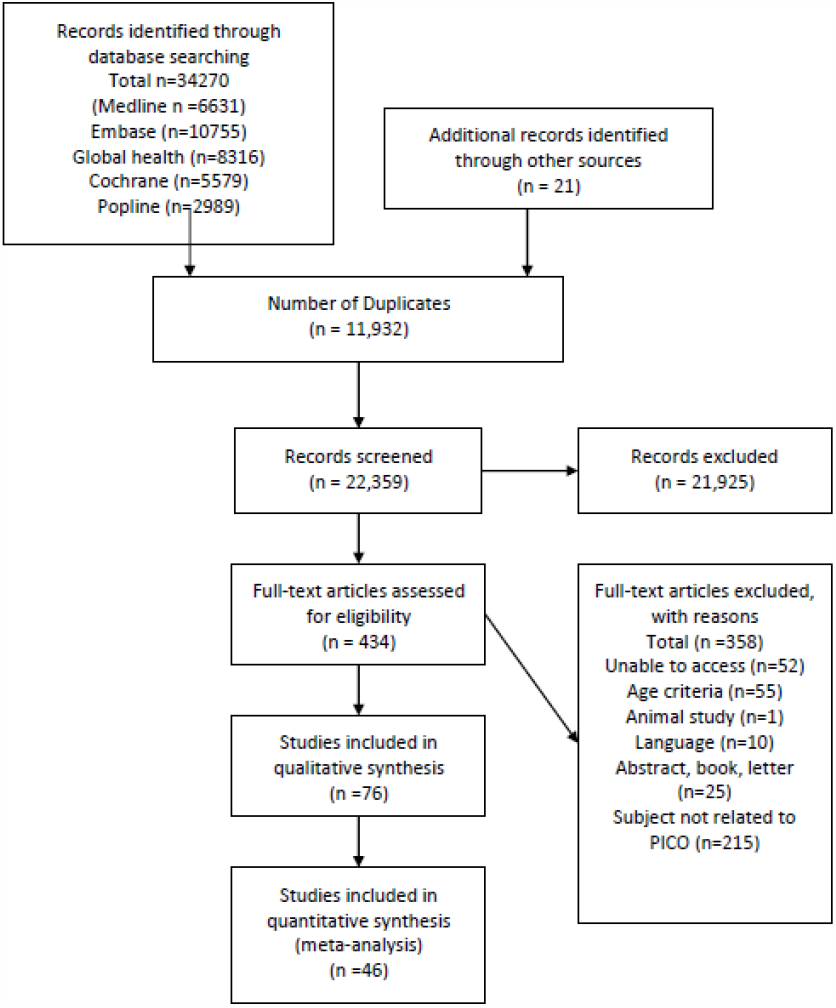
Prisma Flow diagram.

### Study Characteristics

Table 1 shows the characteristics of each of the studies included in the review. The studies selected for the review varied widely in terms of aims and study design. Many were observational, assessing prevalence of undernutrition and related risk factors and many included secondary data analysis. The outcomes, both primary and secondary also varied widely. The studies took place in more than 30 countries (some covered multiple countries). The studies were spread across Central Africa (3/76) East Africa (35/76), East Asia (1/76), North Africa (1/76), Oceania (1/76), South America (2/76), South Asia (10/76), South East Asia (9/76), South West Pacific (1/76), West Africa (8/76) and multiple countries (5/76).

**Table 1.** Study Characteristics table.

| Study | Study design | Country | Region | Sample size | boy/girl difference wasting | boy/girl difference stunting | boy/girl difference underweight | Reasons for differences discussed |
| --- | --- | --- | --- | --- | --- | --- | --- | --- |
| Kismul H, et al. Determinants of childhood stunting in the Democratic Republic of Congo: further analysis of Demographic and Health Survey 2013-14. BMC Public Health. (2018) 18:74 | Cross-sectional | DRC | Central Africa | 8994 | NA | More boys | NA | No |
| Sakisaka K, et al. Nutritional status and associated factors in children aged 0-23 months in Granada, Nicaragua. Public Health. 2006;120(5):400-11. | Cross-sectional | Nicaragua | Central Africa | 755 | NA | More girls | More girls | Yes |
| Vonaesch P, et al. Factors associated with stunting in healthy children aged 5 years and less living in Bangui (RCA). PLoS ONE. 2017;12(8):e0182363. | Cross-sectional | CAR | Central Africa | 414 | NA | More boys | NA | CRC |
| Abdulahi A. et al Nutritional Status of Under Five Children in Ethiopia: A Systematic Review and Meta-Analysis. Ethiop. 2017;27(2):175-88. | Systematic review and meta-analysis | Ethiopia | East Africa | 39585 | Not reported | More boys | Not reported | No |
| Abera L, Dejene T, Laelago T. Prevalence of malnutrition and associated factors in children aged 6-59 months among rural dwellers of damot gale district, south Ethiopia: community based cross sectional study. Intern. 2017;16(111):1-8. | Cross-sectional | Ethiopia | East Africa | 398 | Not reported | NA | More boys | CRC |
| Abraham D, Elifaged H, Berhanu E. Age-related factors influencing the occurrence of undernutrition in north-eastern Ethiopia. BMC Public Health. 2015;15(108). | Cross-sectional | Ethiopia | East Africa | 50 | More boys | More boys | More boys | Yes |
| Altare C. et al. Factors associated with stunting among pre-school children in southern highlands of Tanzania. J Trop Pediatr. 2016;62(5):390-408. | Cross-sectional | Tanzania | East Africa | 3264 | NA | More boys | NA | CRC |
| Asfaw, M. et al Prevalence of undernutrition and associated factors among children aged between six to fifty nine months in Bule Hora district, South Ethiopia. BMC Public Health. 2015;15:41. | Cross-sectional | Ethiopia | East Africa | 796 | More boys | More boys | More boys | Yes |
| Bukusuba J, Kaaya AN, Atukwase A. Predictors of stunting in children aged 6 to 59 months: a case-control study in southwest Uganda. Food and Nutrition Bulletin. 2017;38(4):542-53. | Case control | Uganda | East Africa | 168 | NA | More boys | NA | Yes |
| Chirande, L. et al Determinants of stunting and severe stunting among under-fives in Tanzania: evidence from the 2010 cross-sectional household survey. BMC Pediatr. 2015;15(165). | Cross-sectional | Tanzania | East Africa | 7324 | NA | More boys | NA | Yes |
| Chirwa EW, Ngalawa HPE. Determinants of child nutrition in Malawi. South African Journal of Economics. 2008;76(4):628-40. | Cross-sectional | Malawi | East Africa | 13858 | More boys | More boys | More boys | Yes |
| Condo, J. et al. Sex differences in nutritional status of HIV-exposed children in Rwanda: a longitudinal study. Trop Med Int Health. 2015;20(1):17-23. | Longitudinal | Rwanda | East Africa | 480 | More boys | More boys | More boys | Yes |
| Cruz, L. et al. Factors Associated with Stunting among Children Aged 0 to 59 Months from the Central Region of Mozambique. Nutrients. 2017;9(5):12. | Case control | Mozambique | East Africa | 282 | NA | More boys | NA | Yes |
| Deneke T, et al. Risk factors of underweight in children aged 6-59 months in Ethiopia. Journal of Nutrition and Metabolism. 2017;6368746(33). | Cross-sectional | Ethiopia | East Africa | 642 | NA | NA | More boys | Yes |
| Eskezyiaw A, Tefera C. Predictors of chronic under nutrition (Stunting) among children aged 6-23 months in Kemba Woreda, Southern Ethiopia: a community based cross-sectional study. Journal of Nutrition and Food Sciences. 2015;5(4). | Cross-sectional | Ethiopia | East Africa | 562 | NA | More boys | NA | No |
| Espo M, et al. Determinants of linear growth and predictors of severe stunting during infancy in rural Malawi. Acta Paediatrica. 2002;91(12):1364-70. | Cohort | Malawi | East Africa | 613 | NA | More boys | NA | No |
| Ettyang GA, Sawe CJ. Factors associated with stunting in children under age 2 in the Cambodia and Kenya 2014 Demographic and Health Surveys. Rockville, Maryland, ICF, DHS Program, 2016 Aug.; 2016. | Cross-sectional | Kenya and Cambodia | East Africa | 10,163 | NA | More boys | NA | Yes |
| Fentahun N, Belachew T, Lachat C. Determinants and morbidities of multiple anthropometric deficits in southwest rural Ethiopia. Nutrition. 2016;32(11-12):1243-9. | Longitudinal | Ethiopia | East Africa | 1927 | More girls | More girls | More girls | Yes |
| Geresomo N, et al. Risk factors associated with stunting among infants and young children aged 6-23 | Cross-sectional | Malawi | East Africa | 306 | NA | More boys | NA | Yes |
| months in Dedza district of Central Malawi. African Journal of Food, Agriculture, Nutrition and Development. 2017;17(4):12854-70. |  |  |  |  |  |  |  |  |
| Gewa CA, Yandell N. Undernutrition among Kenyan children: contribution of child, maternal and household factors. Public Health Nutr. 2012;15(6):1029-38. | Cross-sectional | Kenya | East Africa | 3793 | More boys | More boys | More boys | Yes |
| Girmay M, et al. Prevalence and predictors of undernutrition among infants aged six and twelve months in Butajira, Ethiopia: the P-MaMiE Birth Cohort. BMC Public Health. 2010;10(27). | Longitudinal | Ethiopia | East Africa | 1799 | NA | More boys | More boys | Yes |
| Habtom K, et al. Prevalence of acute malnutrition and its associated factors among children aged 6-59 months in Mai-Aini Eritrean refugees' camp, Northern Ethiopia. Journal of Nutrition and Food Sciences. 2015;5(1). | Cross-sectional | Ethiopia | East Africa | 593 | More Boys | NA | More boys | No |
| Haile D, et al. Exploring spatial variations and factors associated with childhood stunting in Ethiopia: Spatial and multilevel analysis. BMC Pediatr. 2016;16 (1) | Cross-sectional | Ethiopia | East Africa | 9893 | NA | More boys | NA | Yes |
| Kinyoki DK, et al. Assessing comorbidity and correlates of wasting and stunting among children in Somalia using cross-sectional household surveys: 2007 to 2010. BMJ Open. 2016;6(3) | Cross-sectional | Somalia | East Africa | 73778 | More boys | More boys | More boys | No |
| Lamirot A, Tariku D, Tariku L. Prevalence of malnutrition and associated factors in children aged 6-59 months among rural dwellers of Damot Gale district, South Ethiopia: community based cross sectional study. Intern. 2017;16(111). | Cross-sectional | Ethiopia | East Africa | 398 | More boys | NA | More boys | CRC |
| Masibo PK, Makoka D. Trends and determinants of undernutrition among young Kenyan children: Kenya Demographic and Health Survey; 1993, 1998, 2003 and 2008-2009. Public Health Nutr. 2012;15(9):1715-27. | Cross-sectional | Kenya | East Africa | 19021 | NA | More boys | More boys | No |
| Matanda DJ, Mittelmark MB, Kigaru DMD. Child undernutrition in Kenya: trend analyses from 1993 to 2008-09. BMC Pediatr. 2014;14(5). | Cross-sectional | Kenya | East Africa | 11938 | More boys | More boys | NA | Yes |
| Medhin G, et al. Prevalence and predictors of undernutrition among infants aged six and twelve | Cohort | Ethiopia | East Africa | 1799 | NA | More boys | More boys | Yes |
| months in Butajira, Ethiopia: the P-MaMiE Birth Cohort. BMC Public Health. 2010;10:27. |  |  |  |  |  |  |  |  |
| Mgongo M, et al. Underweight, Stunting and Wasting among Children in Kilimanjaro Region, Tanzania; a Population-Based Cross-Sectional Study. International Journal of Environmental Research and Public Health. 2017;14(5). | Cross-sectional | Tanzania | East Africa | 1870 | more boys | more boys | More boys | CRC |
| Moges B, et al. Magnitude of stunting and associated factors among 6-59 months old children in Hossana Town, Southern Ethiopia. Journal of Clinical Research and Bioethics. 2015;6(1). | Cross-sectional | Ethiopia | East Africa | 734 | NA | More boys | NA | No |
| Ndemwa M, et al. Nutritional status and association of demographic characteristics with malnutrition among children less than 24 months in Kwale County, Kenya. Pan Afr Med J. 2017;28:265. | Cross-sectional | Kenya | East Africa | 380 | NA | More boys | More boys | Yes |
| Ndiku M, et al. Gender inequality in food intake and nutritional status of children under 5 years old in rural Eastern Kenya. Eur J Clin Nutr. 2011;65(1):26-31. | Cross-sectional | Kenya | East Africa | 629 | More girls | More girls | More girls | Yes |
| Ntenda PAM, Chuang YC. Analysis of individual-level and community-level effects on childhood undernutrition in Malawi. Pediatrics and Neonatology. 2017. | Cross-sectional | Malawi | East Africa | 6384 | More boys | More boys | More boys | Yes |
| Olwedo MA, et al. Factors associated with malnutrition among children in internally displaced person's camps, northern Uganda. Afr Health Sci. 2008;8(4):244-52. | Cross-sectional | Uganda | East Africa | 672 | More boys | More boys | NA | Yes |
| Rakotomanana H, et al. The determinants of stunting in children under five in madagascar. FASEB Journal Conference: Experimental Biology. 2016;30(Meeting Abstracts). | Cross-sectional | Madagascar | East Africa | 4774 | NA | More boys | NA | Yes |
| Tadesse AW, et al. Comparison of mid-upper arm circumference and weight-for-height to diagnose severe acute malnutrition: a study in Southern Ethiopia. Nutrients. 2017;9(3). | Cross-sectional | Ethiopia | East Africa | 622 | NA | More boys | NA | Yes |
| Yisak H, Gobena T, Mesfin F. Prevalence and risk factors for under nutrition among children under five at Haramaya district, Eastern Ethiopia. BMC Pediatr. 2015;15:212. | Cross-sectional | Ethiopia | East Africa | 791 | More boys | More boys | More boys | No |
| Yourkavitch J. Trends and inequalities in young child nutrition in Rwanda: further analysis of the 2014-15 demographic and health survey: DHS Further Analysis Report; 2018. | Cross-sectional | Rwanda | East Africa | 1572 | More boys | More boys | More boys | Yes |
| Jiang Y, et al. Prevalence and risk factors for stunting and severe stunting among children under three years old in mid-western rural areas of China. Child Care Health Dev. 2015;41(1):45-51. | Cross-sectional | china | East Asia | 1115 | NA | More boys | NA | CRC |
| Díez-Navarro A, et al. Female eco-stability and severe malnutrition in children: Evidence from humanitarian aid interventions of Action Against Hunger in African, Asian and Latin American countries. Nutricion Clinica y Dietetica Hospitalaria. 2017;34:127-34. | Cross-sectional | Multiple (Africa, Latin America, Asia) | Multiple | 367258 | More boys | More boys | More boys | Yes |
| Khara T, et al. Children concurrently wasted and stunted: A meta-analysis of prevalence data of children 6-59 months from 84 countries. Maternal and Child Nutrition. 2018;14 (2) | Cross-sectional | Multiple (global) | Multiple | 570930 | More boys | More boys | NA | Yes |
| Keino S, et al. Determinants of stunting and overweight among young children and adolescents in sub-Saharan Africa. Food and Nutrition Bulletin. 2014;35(2):167-78. | Systematic review | multiple (sub-Saharan Africa) | Multiple |  | NA | More boys | NA | No |
| Myatt M, et al. Children who are both wasted and stunted are also underweight and have a high risk of death: a descriptive epidemiology of multiple anthropometric deficits using data from 51 countries. Arch Public Health. 2018;76:28. | Meta-analysis | Multiple (global) | Multiple | 1,796,991 | More boys reported with concurrent wasting and stunting | NA |  | Yes |
| Wamani H, et al. Boys are more stunted than girls in sub-Saharan Africa: a meta-analysis of 16 demographic and health surveys. BMC Pediatr. 2007;7:17. | Cross-sectional | Multiple (sub-Saharan Africa) | Multiple | 64000 | NA | More boys | NA | Yes |
| El-Taguri A, et al. Risk factors for stunting among under-fives in Libya. Public Health Nutr. 2009;12(8):1141-9. | Cross-sectional | Libya | North Africa | 4498 | NA | More boys | NA | No |
| Choy, C. et al. Child, maternal and household-level correlates of nutritional status: a cross-sectional study among young Samoan children. Public Health Nutr. 2017;20(7):1235-47. | Cross-sectional | Samoa | Oceania | 305 | NA | More boys | NA | Yes |
| Castro B.A. et al. Factors associated with stunted growth in children below 11 years of age in Antioquia, Colombia, 2004. Colombia Medica. 2011;42(2):207-14. | Cross-sectional | Columbia | South America | 2967 under 5's | NA | More girls | NA | Yes |
| Correia, L. et al. Prevalence and determinants of child undernutrition and stunting in semiarid region of Brazil. Rev Saude Publica. 2014;48(1):19-28. | Cross-sectional | Brazil | South America | 6046 | More girls | More girls | NA | No |
| Aguayo VM, Badgaiyan N, Paintal K. Determinants of child stunting in the Royal Kingdom of Bhutan: an in-depth analysis of nationally representative data. Maternal and Child Nutrition. 2015;11(3):333-45. | Cross-sectional | Bhutan | South Asia | 2085 | NA | More boys | NA | CRC |
| Aguayo VM, Badgaiyan N, Dzied L. Determinants of child wasting in Bhutan. Insights from nationally representative data. Public Health Nutr. 2017;20(2):315-24. | Cross-sectional | Bhutan | South Asia | 2028 | more boys | NA | NA | No |
| Baig-Ansari, N. et al. Child's gender and household food insecurity are associated with stunting among young Pakistani children residing in urban squatter settlements. Food Nutr Bull. 2006;27(2):114-27. | Cross-sectional | Pakistan | South Asia | 399 | NA | More girls | NA | Yes |
| Biswas S, Bose K. Sex differences in the effect of birth order and parents' educational status on stunting: a study on Bengalee preschool children from eastern India. Homo. 2010;61(4):271-6. | Cross-sectional | India | South Asia | 161 | More boys | More girls | More boys | Yes |
| Gupta A. Assessing stunting and predisposing factors among children. Asian Journal of Pharmaceutical and Clinical Research. 2017;10(10):364-71. | Cross-sectional | India | South Asia | 440 | NA | More girls | NA | Yes |
| Khan, A.T. et al. Prevalence and associated factors of malnutrition among children under-five years in Sindh, Pakistan: a cross-sectional study. Tropical Medicine and International Health. 2016;22 (Supplement 1):317-8. | Cross-sectional | Pakistan | South Asia | 3964 | More boys | More boys | More boys | No |
| Kumar D, et al. Socio-demographic factors affecting the nutritional status of the under three children in Chandigarh, UT. Healthline, Journal of Indian Association of Preventive and Social Medicine. 2015;6(1):46-52. | Cross-sectional | India | South Asia | 424 | NA | NA | More girls | Yes |
| Sand A, et al. Determinants of severe acute malnutrition among children under five years in a | Cross-sectional | Pakistan | South Asia | 105 | More boys | NA | NA | No |
| rural remote setting: A hospital based study from district Tharparkar-Sindh, Pakistan. Pakistan Journal of Medical Sciences. 2018;34(2):260-5. |  |  |  |  |  |  |  |  |
| Shaikh S, et al. Lean body mass in preschool aged urban children in India: gender difference. Eur J Clin Nutr. 2003;57(3):389-93. | Cross-sectional | India | South Asia | 245 | More boys | More girls | More girls | Yes |
| Shashank KJ, Angadi MM. Gender disparity in health and nutritional status among under-five children in a rural field practice area of Shri BM Patil Medical College. International Journal of Medical Science and Public Health. 2016;5(2):217-20. | Cross-sectional | India | South Asia | 161 | More boys | More girls | More boys | Yes |
| Adair LS, Guilkey DK. Age-specific determinants of stunting in Filipino children. J Nutr. 1997;127(2):314-20. | Longitudinal | Philippines | South East Asia | 3080 | NA | More boys | NA | Yes |
| Ahmed A.M. et al Determinants of undernutrition in children under 2 years of age from rural Bangladesh. Indian Pediatr. 2012;49(10):821-4. | Cross-sectional | Bangladesh | South East Asia | 8858 | more boys | More boys | More boys | CRC |
| Choudhury, N. et al. Determinants of age-specific undernutrition in children aged less than 2 years-the Bangladesh context. Matern Child Nutr. 2017;13(3). | Surveillance | Bangladesh | South East Asia | 10291 | More boys | More boys | More boys | CRC |
| Chowdhury, M.R. et al. Risk Factors for Child Malnutrition in Bangladesh: A Multilevel Analysis of a Nationwide Population-Based Survey. J Pediatr. 2016;172:194-201.e1. | Cross-sectional | Bangladesh | South East Asia | 7568 | No difference found | More girls | More girls | CRC |
| Dancer D, Rammohan A, Smith MD. Infant mortality and child nutrition in Bangladesh. Health Econ. 2008;17(9):1015-35. | Cross-sectional | Bangladesh | South East Asia | 5172 | More girls | More girls | NA | Yes |
| Islam MM, et al. Risk factors of stunting among children living in an urban slum of Bangladesh: findings of a prospective cohort study. BMC Public Health. 2018;18(1):197. | Cohort | Bangladesh | South East Asia | 265 | NA | More boys | NA | Yes |
| Khambalia A, et al. Prevalence and sociodemographic factors of malnutrition among children in Malaysia. Food Nutr Bull. 2012;33(1):31-42. | Literature review | Malaysia | South East Asia | NA | NA | More boys | No | NA |
| Phengxay M, et al. Risk factors for protein-energy malnutrition in children under 5 years: study from Luangprabang province, Laos. Pediatr Int. 2007;49(2):260-5. | Cross-sectional | Laos | South East Asia | 798 | More girls | More boys | More boys | No |
| Ramli, A. et al. Prevalence and risk factors for stunting and severe stunting among under-fives in North Maluku province of Indonesia. BMC Pediatr. 2009;9:64. | Cross-sectional | Indonesia | South East Asia | 2168 | NA | More boys | NA | No |
| Olita'a D, et al. Risk factors for malnutrition in children at Port Moresby General Hospital, Papua New Guinea: A case-control study. J Trop Pediatr. 2014;60(6):442-8. | Case control | Papua New Guinea | South West Pacific | 50 | More girls | More girls | More girls | Yes |
| Akombi BJ, et al. Stunting and severe stunting among children under-5 years in Nigeria: A multilevel analysis. BMC Pediatr. 2017;17(1):15. | Cross-sectional | Nigeria | West Africa | 24529 | NA | More boys | NA | Yes |
| Amugsi DA, Mittelmark MB, Lartey A. An analysis of socio-demographic patterns in child malnutrition trends using Ghana demographic and health survey data in the period 1993-2008. BMC Public Health. 2013;13:960. | Cross-sectional | Ghana | West Africa | 7235 | More boys | More boys | More boys | No |
| Bork KA, Diallo A. Boys Are More Stunted than Girls from Early Infancy to 3 Years of Age in Rural Senegal. J Nutr. 2017;147(5):940-7. | Cohort | Senegal | West Africa | 7319 | More boys | More boys | NA | Yes |
| Darteh EK, Acquah E, Kumi-Kyereme A. Correlates of stunting among children in Ghana. BMC Public Health. 2014;14:504 | Cross-sectional | Ghana | West Africa | 2379 | NA | More boys | NA | No |
| Garenne M, et al. Concurrent wasting and stunting among under-five children in Niakhar, Senegal. Matern Child Nutr. 2018:e12736. | Longitudinal | Senegal | West Africa |  | More boys | More boys | NA | Yes |
| Miah RW, Apanga PA, Abdul-Haq Z. Risk factors for undernutrition in children under five years old: evidence from the 2011 Ghana Multiple Indicator Cluster Survey. Journal of AIDS and Clinical Research. 2016;7(7). | Cross-sectional | Ghana | West Africa | 7550 | more boys | more boys | more boys | Yes |
| Olusanya BO, Wirz SL, Renner JK. Prevalence, pattern and risk factors for undernutrition in early infancy using the WHO Multicentre Growth Reference: a community-based study. Paediatr Perinat Epidemiol. 2010;24(6):572-83. | Cross-sectional | Nigeria | West Africa | 5994 | More boys | more boys | More boys | No |
| Poda GG, Hsu C, Chao JCJ. Factors associated with malnutrition among children | Cross-sectional | Burkina Faso | West Africa | 6337 | More boys | More boys | More boys | CRC |

Where sample size was clearly stated, the included studies involved 3,133,992 participants. Distribution of boys and girls was not provided for all studies but, where they were, results showed a total of 1,490,691 (47.6%) girls and 1,532,951 (48.9%) boys. Inclusion criteria for age entailed a mix of studies covering all children aged 0-59 months with others focused on subsets of these children.

### Meta-analysis

We identified 76 studies that had measured undernutrition in the form of wasting, stunting and underweight and reviewed them for inclusion in the meta-analysis. 46 studies included extractable data, fully disaggregated by sex and were therefore eligible for inclusion, 42 of these were cross sectional and 4 were longitudinal (in which case the most recent prevalence data were used). Results from the analysis are presented in the forest plots in figure 2.

**Figure 2.**
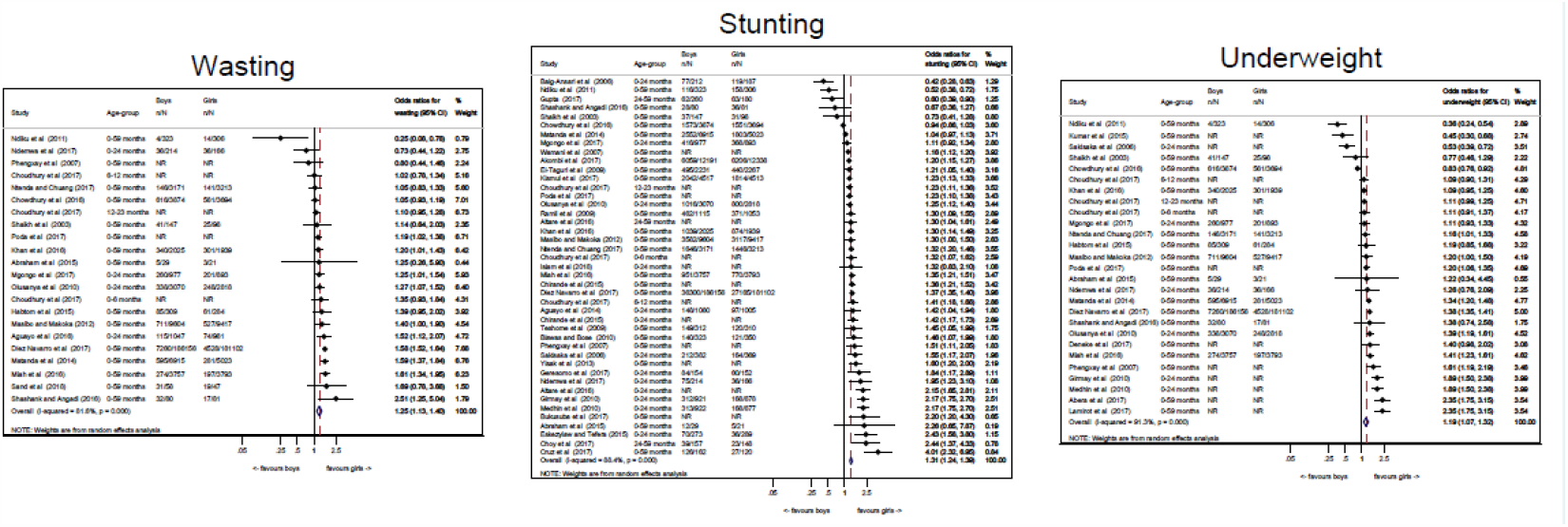
Forest plots showing pooled analysis for wasting, stunting and underweight.

### Pooled analysis by outcome

20 studies were included in the pooled analysis of wasting. In 17 of the 20 studies, wasting was more prevalent in boys than girls, with a statistically significant difference in 11/17 of the studies. In the remaining 3 studies, wasting was more prevalent in girls than boys, with a significant difference in 1/3 of the studies. The pooled results of individual studies for wasting showed that boys had 26% higher odds of being wasted than girls (pooled OR 1.26, 95%CI 1.13-1.40, p<0.001).

39 studies were included in the pooled analysis of stunting. In 33 of the 39 studies, stunting was more prevalent in boys than girls, with a statistically significant difference in 29/33 of the studies. In the remaining 6 studies, stunting was more prevalent in girls than boys, with a significant difference in 3/6 of the studies. The pooled results for stunting showed that boys had 31% higher odds of being stunted than girls (pooled OR 1.31 95%CI 1.24-1.39, p<0.001).

25 studies were included in the pooled analysis of underweight. In 20 of the 25 studies, underweight was more prevalent in boys than in girls, with a statistically significant difference in 12/20 of the studies. In the remaining 5 studies, girls were more likely to be underweight than boys, with a significant difference in 4/5 of the studies. The pooled results for underweight showed that boys had 19% higher odds of being underweight than girls (OR 1.19, 95%CI 1.07-1.32, p=0.001).

### Pooled analysis by region

When organised by region, the odds of boys being malnourished were nearly always higher than for girls for wasting, stunting and underweight. For wasting, the odds were higher for boys than for girls in all regions. For stunting, the odds were higher for boys than for girls in all regions except South Asia (pooled OR 0.88, 95%CI 0.62-1.26 p=0.492), where there was no difference by sex. For underweight, the odds were higher for boys than for girls in all regions except Central America (OR 0.53, 95%CI 0.40-0.72, p=0.000), although this finding was from a single study, and South Asia (pooled OR 0.84, 95%CI 0.52-1.35, p=0.475). Results from the analysis are presented in table 2.

**Table 2.** Odds of boys being undernourished compared with girls by regions and age groups.

| Region/<br>Age groups | No. of<br>studies<br>of<br>Wasting | Pooled OR<br>(95% CI) | P Value | No. of<br>studies<br>of<br>stunting | Pooled OR<br>(95% CI) | P Value | No. of<br>studies of<br>underweight | Pooled OR<br>(95% CI) | P Value |
| --- | --- | --- | --- | --- | --- | --- | --- | --- | --- |
| **Africa** |  |  |  |  |  |  |  |  |  |
| East | 8 | 1.18<br>(0.95-1.47) | 0.126 | 17 | 1.53<br>(1.33-1.77) | <0.001 | 13 | 1.35<br>(1.12-1.63) | 0.002 |
| West | 3 | 1.34<br>(1.12-1.59) | 0.001 | 4 | 1.24<br>(1.18-1.30) | <0.001 | 3 | 1.32<br>(1.19-1.47) | <0.001 |
| Central |  |  |  | 1 | 1.23<br>(1.13-1.33) | <0.001 |  |  |  |
| North |  |  |  | 1 | 1.21<br>(1.05-1.40) | 0.009 |  |  |  |
| **Oceania** |  |  |  | 1 | 2.44<br>(1.37-4.33) | 0.002 |  |  |  |
| **Asia** |  |  |  |  |  |  |  |  |  |
| South | 5 | 1.39<br>(1.12-1.72) | 0.003 | 7 | 0.88<br>(0.62-1.26) | 0.492 | 4 | 0.84<br>(0.52-1.35) | 0.475 |
| South East | 3 | 1.08<br>(0.99-1.17) | 0.092 | 5 | 1.25<br>(1.08-1.45) | 0.003 | 3 | 1.09<br>(0.91-1.32) | 0.350 |
| **Central**<br>**America** |  |  |  | 1 | 1.56<br>(1.17-2.07) | 0.003 | 1 | 0.53<br>(0.40-0.72) | <0.001 |
| **Multiple**<br>**studies** | 1 | 1.58<br>(1.52-1.64) | <0.001 | 2 | 1.26<br>(1.07-1.49) | 0.006 | 1 | 1.38<br>(1.35-1.41) | <0.001 |
| **Age group** |  |  |  |  |  |  |  |  |  |
| 0-24m | 5 | 1.19<br>(1.06-1.34) | 0.004 | 14 | 1.47<br>(1.27-1.70) | <0.001 | 7 | 1.20<br>(0.99-1.46) | 0.060 |
| 24-59m |  |  |  | 3 | 1.21<br>(0.63-2.33) | 0.572 |  |  |  |
| 0-59m | 15 | 1.30<br>(1.13-1.48) | <0.001 | 24 | 1.24<br>(1.16-1.32) | <0.001 | 18 | 1.18<br>(1.03-1.34) | 0.014 |

### Pooled analysis by age

When organised by age groups, the odds of boys being wasted, stunted or underweight were higher in all age categories for boys than for girls. Results from the analysis are presented in table 2.

We repeated the analysis for relative risk and found the results were consistent with results for odds ratios. For wasting, stunting and underweight, there was strong evidence of between-study heterogeneity of effect, which meta-regression by age group and region did not explain.

### Risk of bias within studies

The quality assessment is presented in table 3. All studies presented appeared to be of acceptable quality. It is worth noting however that a number of studies were excluded prior to this process due to the absence of suitable data. The main differences in quality were in the domain which assessed whether sex differences were acknowledged and explored (see qualitative synthesis section).

**Table 3:**
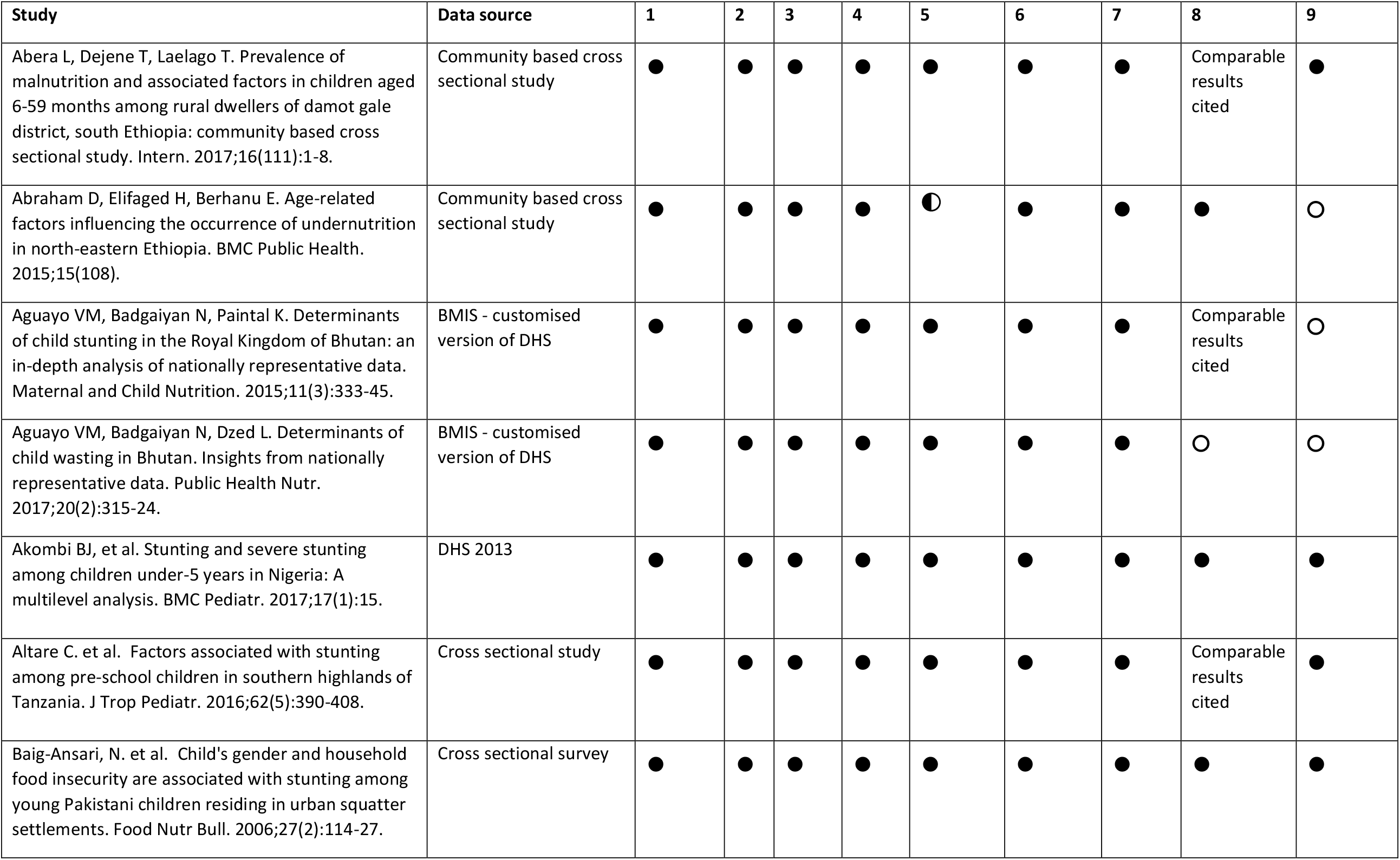
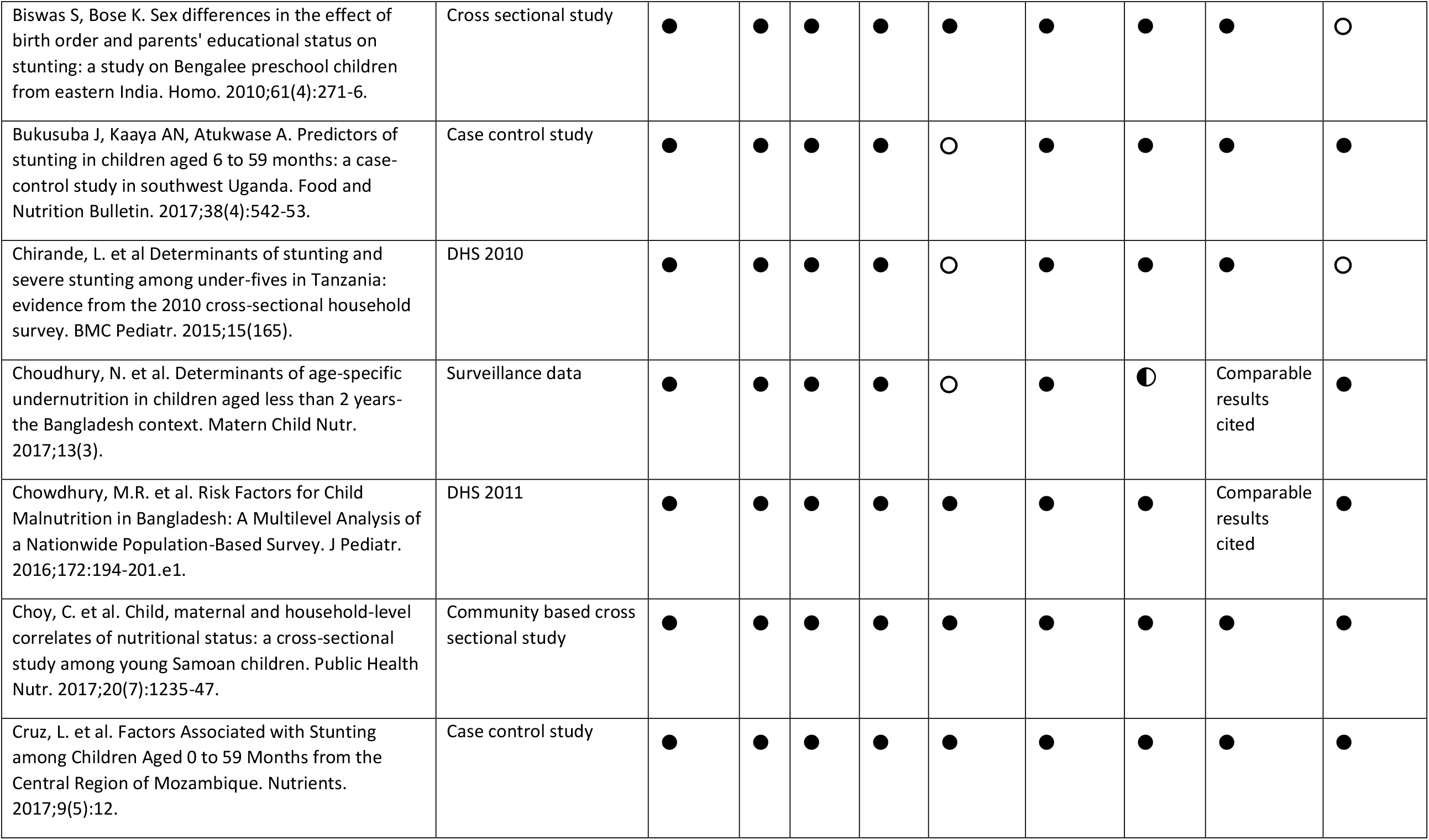
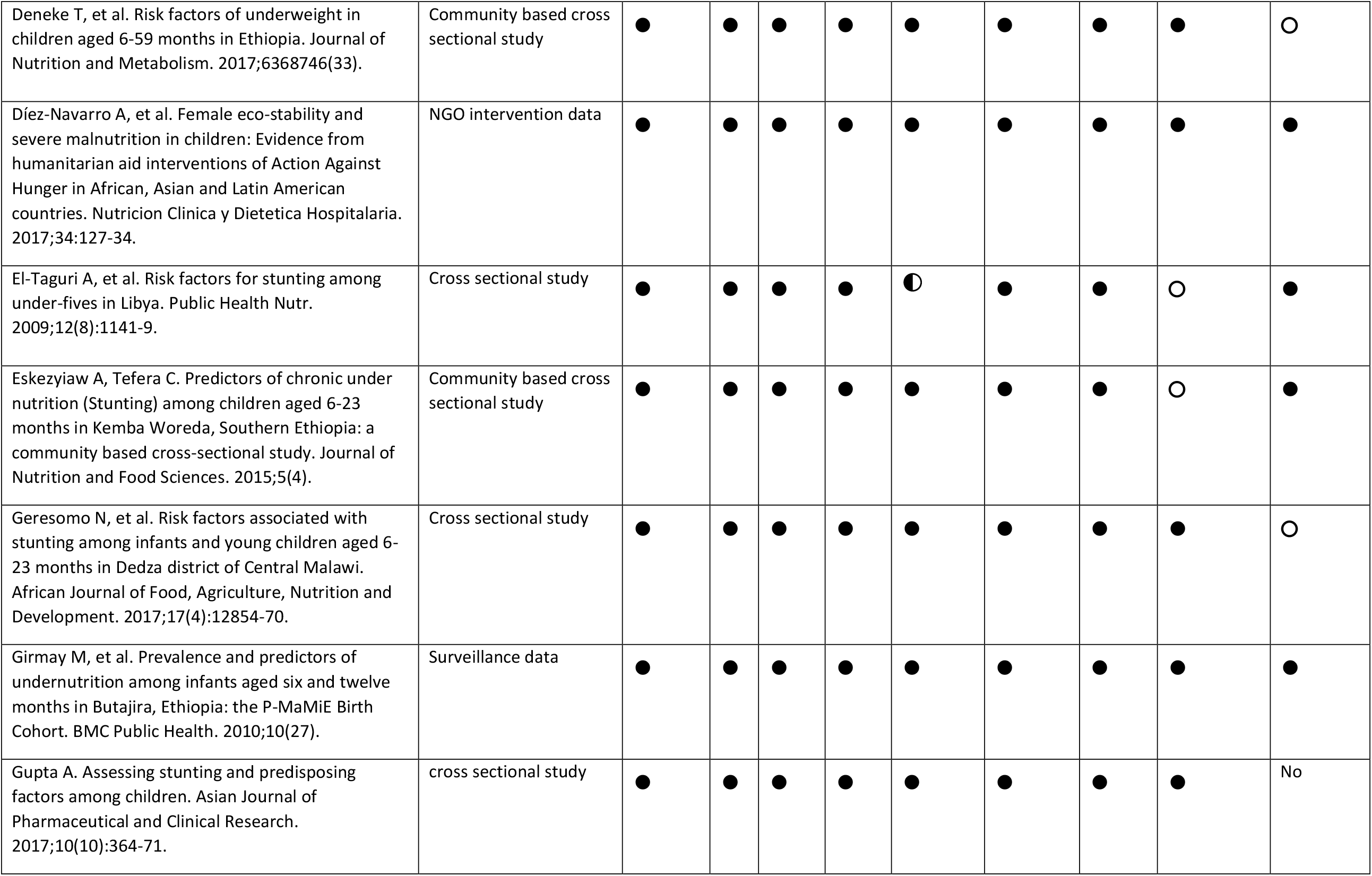
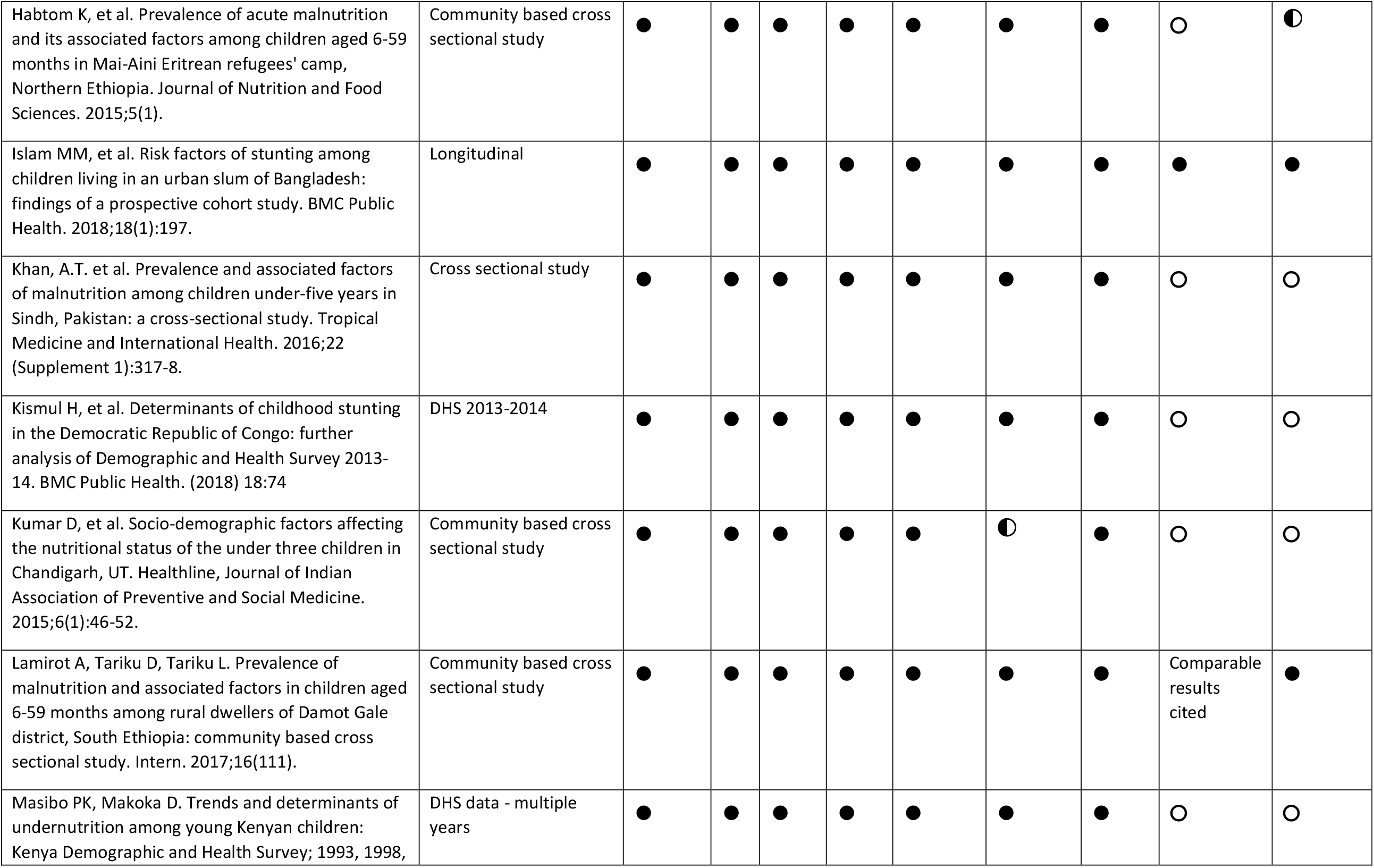
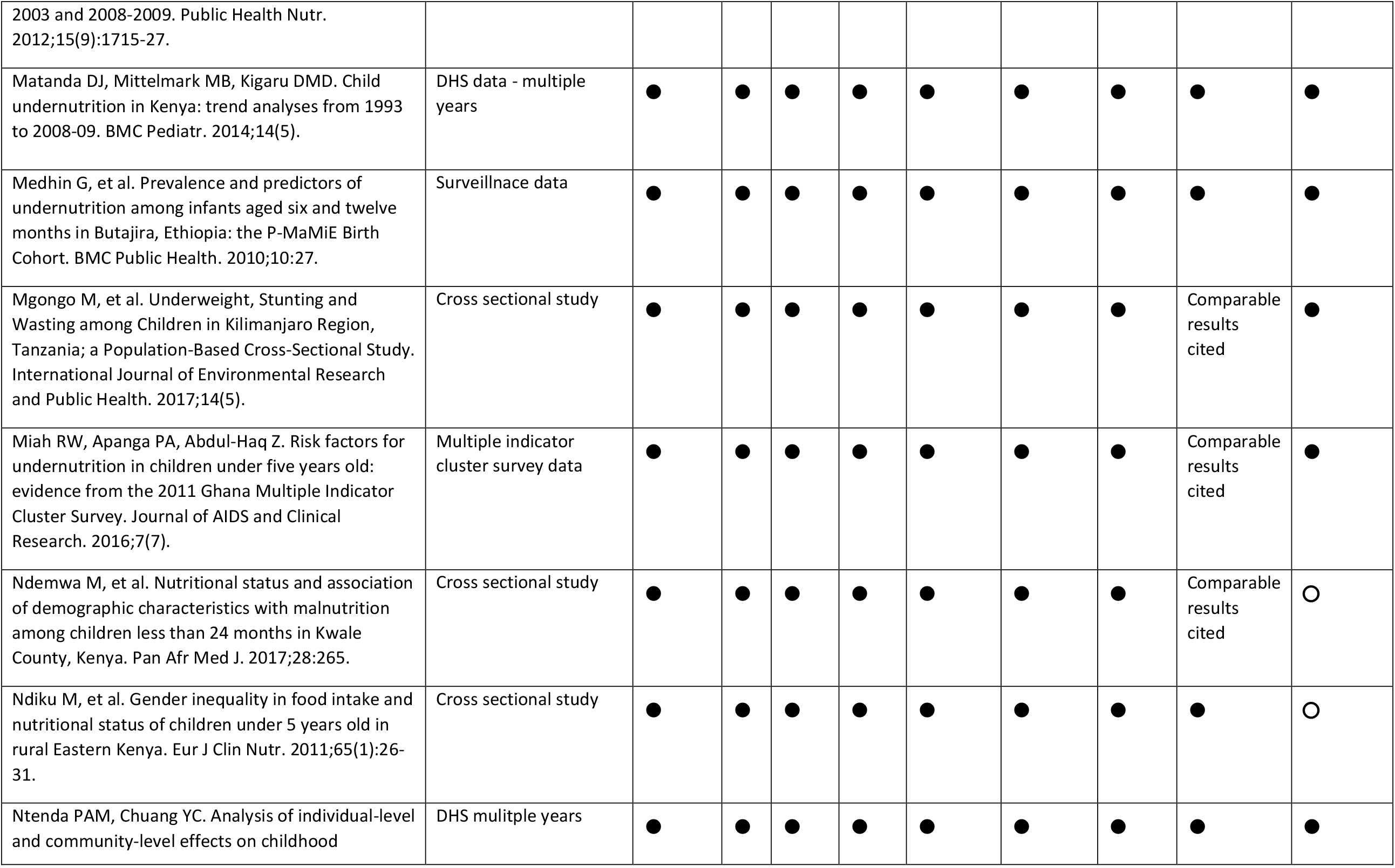
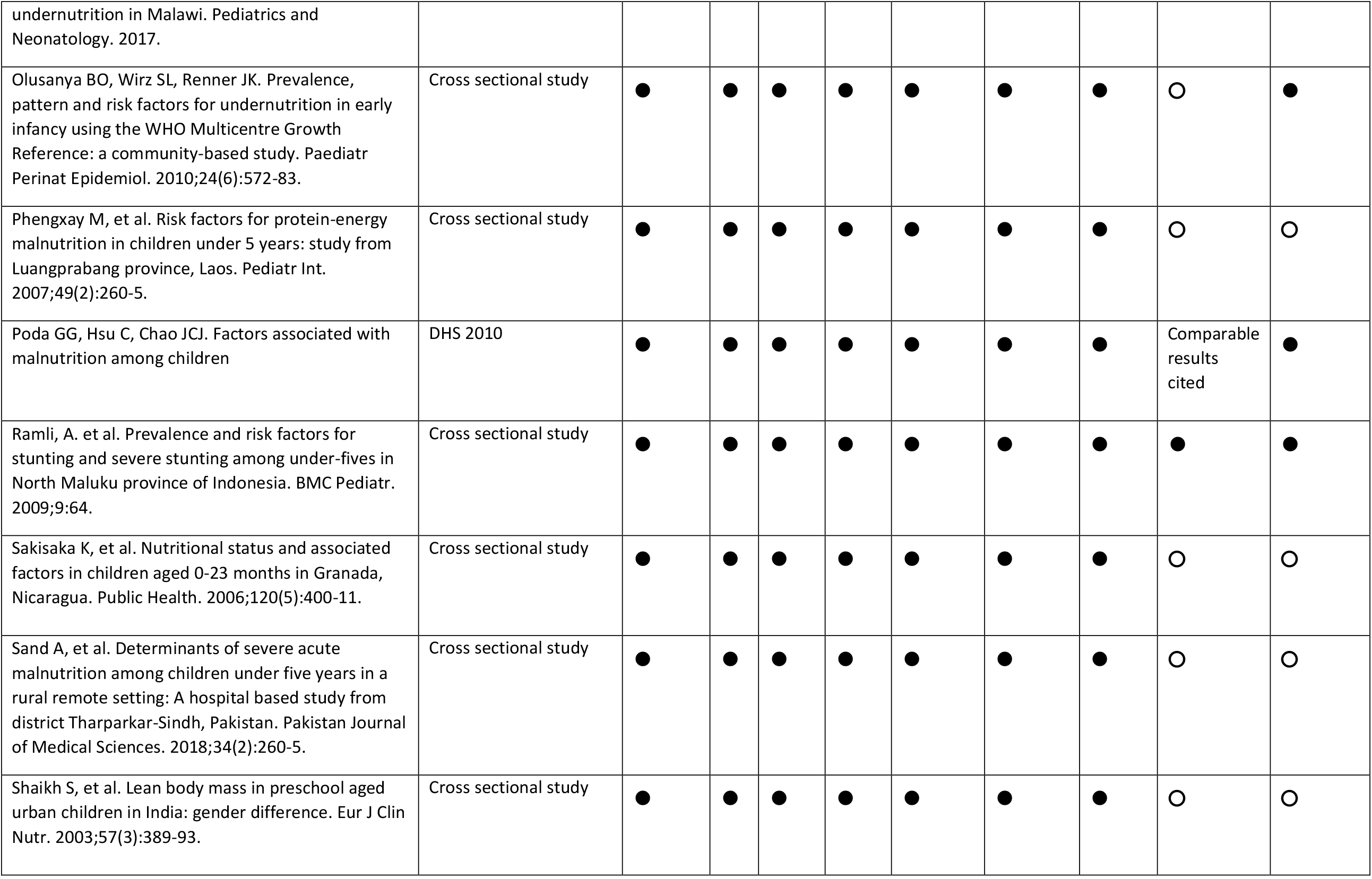
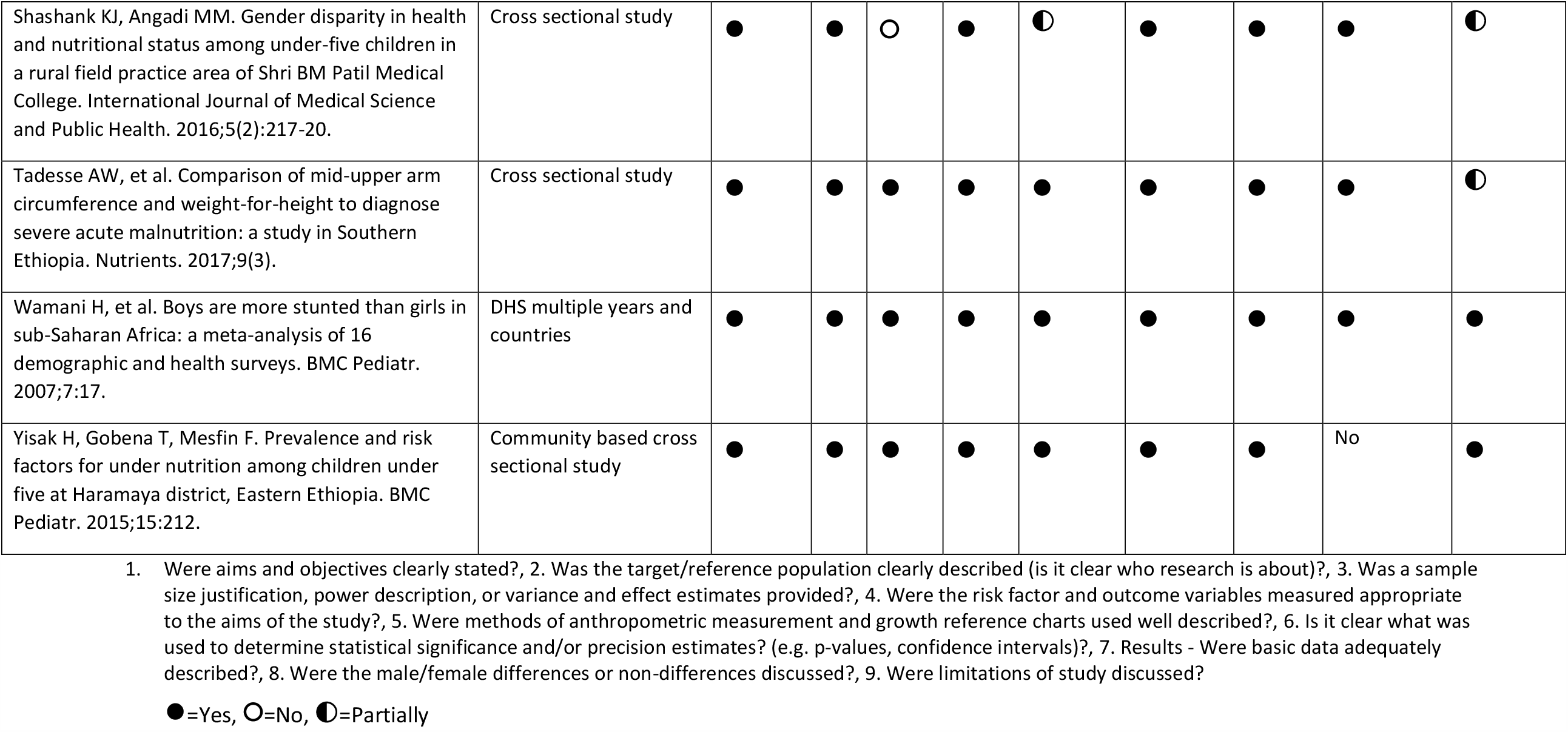
Risk of Bias assessment.

### Qualitative synthesis

76 studies reported on outcomes related to undernutrition – wasting, stunting and underweight. From this, 38/76 studies reported on wasting as an outcome with 31/38 (81%) reporting a higher prevalence of wasting in boys, 6/38 (16%) reporting a higher prevalence of wasting in girls, 1/38 (3%) reporting no difference in the prevalence of wasting between boys and girls. 68/76 studies reported on stunting as an outcome. 55/68 (81%) reported a higher prevalence of stunting in boys and 13/68 (19%) reported higher prevalence of stunting in girls. 37/76 studies reported on underweight as an outcome. 30/37 (81%) reported higher prevalence of being underweight in boys, 7/37 (19%) reported a higher prevalence of underweight in girls.

We reviewed the discussion sections of the reports to see if these findings were explicitly acknowledged and if explanations were offered. 44/76 (58%) of the studies discussed the findings, 11/76 (14%) studies cited articles with similar findings but did not speculate as to the causes of these differences and 21/76 (28%) of the studies did not discuss the findings related to sex differences at all.

Among those study reports that did offer explanations for sex differences the reasons varied widely and were often conjectural. We coded explanations as either biological (6/44; 14%), social (21/44; 48%), or a combination of the two (17/44; 38%). Biological reasons varied from a simple statement of “biological differences” to more detailed exploration of sex differences in the immune and endocrine system between boys and girls. Social reasons given varied widely and were almost entirely conjectural, with exceptions identified through regression analysis related to son preference and related to sibling order and sex. Other social reasons given were gender dynamics, preferential feeding practices for either boys or girls, infant and young child feeding practices such as early weaning for boys and children’s behaviours whereby girls might stay closer to the home and have more access to food being cooked whilst boys play outside and in turn eat less while expending more energy.

## Discussion

This review offers a systematic look at sex differences over a wide geographical area. The studies included in the meta-analysis show that boys aged 0-59 months are much more likely to be wasted, stunted and underweight using anthropometric case definitions than girls. This indicates sex differences in susceptibility to undernutrition. The reasons currently provided for these differences vary and are often speculative rather than informed by direct evidence.

When stratified by region, the results also showed that boys are more likely to be wasted, stunted, or underweight than girls. There were however some exceptions where odds ratios were reduced or reversed for boys with respect to undernutrition, in East Africa, Central America, South and South-East Asia. The differences in Central America were based solely on one study, with a limited sample size and therefore need to be interpreted with caution. Our analysis potentially masks some of the complexities of regional variations in sex differences, particularly in South and South East Asia as many studies from these regions did not qualify for inclusion in the meta-analysis due to insufficient data. It is possible these differences might be under or over-estimated. In reviewing the individual studies identified in the main search, results from this region are inconsistent and often conflicting compared with those coming from other regions of the world, such as Africa, which show a more consistent pattern of male disadvantage, a finding resonating with other studies. [5, 21] The inconsistencies in findings for parts of South and South-East Asia, however, may be explained in part by well-described social preferences for males [22], and warrant further investigation. Such differences have also been described for under-five mortality, with excess female child mortality for certain diseases, and according to socio-economic status, birth order and family composition. [23-26]

These findings challenge commonly held assumptions within the nutrition community that girls are more likely to be affected by undernutrition. Recent studies focused on the relationship between wasting and stunting have also highlighted similar findings showing boys are more likely to be concurrently wasted and stunted than girls [4, 27-29] and have identified this as an unexpected finding.

We found that even where sex differences are reported, they are not always acknowledged or explored. Just over a quarter of studies (28%) did not provide any discussion on reported differences and 15% cited similar findings but did not consider causes. Where explanations for sex differences in the prevalence of undernutrition were offered, nearly half (48%) of the studies reviewed offered explanations related to social reasons or based on speculation or pre-conceived supposition rather than evidence. The search criteria used (which filtered articles to those which use terms related to sex or gender in the abstract) might have introduced some bias here with a potential over-estimation of studies that report and explore the issue of sex differences.

When stratified by age, the meta-analysis also shows that boys are at higher risk across all age groups, though again, our analysis potentially masks some of the complexities in age as detailed analysis of different age groups was not possible. Whilst the results for age show that boys are more likely to be stunted than girls, the odds ratios are lower in the older age group compared with the younger group. Limited data in the 24-59-month age category, especially for wasting and underweight however mean results must be interpreted with caution. These tentative results might indicate any sex-specific risks differ at different ages: further study is warranted. Two studies exploring concurrent wasting and stunting [28, 29] found it to be a condition that affects children below 30 months more than it does older children, and found that sex ratios in undernourished children change with age, with a higher susceptibility for boys up to 30 months that then disappeared. Alongside other studies, [30] they suggest that sex hormones, specifically testosterone, luteinizing hormone and follicle stimulating hormones might play a role in this. Selection effects might also contribute to this, whereby if boys are more likely to die than girls, the remaining pool of boys would represent healthy survivors.

Adair and Guilkey [31] studied children in the Philippines and found males were more likely to become stunted in the first year of life (using the NCHS reference), but females were more likely than males to become stunted in the second year. They suggest differences in parental care-giving behaviours may partly account for this finding, but these were not measured in the study. Bork and Diallo [32] also found evidence of interaction between age and sex in that the deficit in boys compared with that in girls increased between the first and second years of life, regardless of the indicator used. The differences in height status were however sensitive to the growth reference chosen; they were greater when assessed using the 2006 WHO growth standards than when using the NCHS growth reference.

Sex differences in undernutrition may vary not only by geographical area, but also over time. When diseases causing undernutrition known to be more severe among girls, such as measles, whooping cough and tuberculosis, disappear because of vaccination, lower transmission and better feeding, the disadvantage of boys might increase. Conversely, if efficient nutrition programmes are conducted, the disadvantage of boys might be reduced over the years.

Interpretation of these findings into implications for practice and policy are limited at this stage but do warrant consideration and some degree of change. As a minimum, the systematic collection and reporting of disaggregated data by age and sex should be included in the design of programmes and assessments in all settings. Where differences are observed, particularly in programme admissions, these should be interpreted in light of sex differences in population burden in order to draw conclusions as to whether programmes are proving equally accessible to boys and girls, and then the potential causes of these differences should be considered and/or investigated. At present, boys’ vulnerability to undernutrition is rarely a consideration in the design of nutrition programming, nor the formulation of policy. Moreover, some high-level international nutrition policies explicitly focus on the vulnerability of women and girls (e.g. The Scaling Up Nutrition Movement Road Map for 2016–2020, cited Khara et al 2017). Similarly, the recent IASC guidance on gender in humanitarian action [33] recognises the inequity in food intake that may be faced by women and girls in crises but makes no reference to higher levels of undernutrition amongst boys. The absence of any reflection on gender, or the misuse of the term to highlight solely the health of women and girls, is likely to unintentionally reinforce inequalities in health. [7] In the Nutrition for Growth 2020 summit (https://nutritionforgrowth.org/) and beyond, a major focus will be on inequities in undernutrition and how they affect different groups in different locations. The emerging findings from this review have significance in ensuring consideration of these sex differences through an equity lens.

### Strengths and limitations

One of the strengths of this study lies in the systematic approach that was chosen and its primary objective to review sex differences in undernutrition over a wide geographical area. However, there are areas where bias has potentially been introduced.

Firstly, screening for studies to be included in this study was conducted by only one of the authors. Whilst we employed systems to ensure contentious articles were discussed among two or more authors, we recognise that not using double screening is a limitation [34].

Secondly, the search strategy looking for explicit mention of sex or gender in the abstract might have biased towards studies that reported on sex and gender in the abstract, or towards studies that found a significant difference, and therefore sex differences might be under or over-reported in this study. Likewise, the search may have limited the analysis as there are potentially missed studies which include sex as a variable in analysis but without focusing on mention of sex in the study abstracts. Similarly, there may be a degree of publication bias whereby sex differences are simply not considered or reported.

The search criteria also encompassed a large number of studies with differing objectives meaning a limited degree of homogeneity. Few studies directly assessed the true relation between sex and undernutrition. This analysis is therefore potentially biased by healthy survivors – those children that have survived to be included in studies. We do not believe however that our results would be significantly different considering the evidence presented on male vulnerability. Though this analysis included some secondary DHS data, the subject in question could benefit from a systematic analysis of DHS, MICS and or nutrition survey data. Though it is not believed that the outcome of the odds ratios of sex differences would be different, further analysis might help improve understanding of some of the complexities of age, context, dual burdens of undernutrition and sex differences and the implications for programmers.

The rigour of findings of the analysis are limited in relation to age as the grouping and degree of available data potentially masks some of the differences at stages of the lifecycle, similarly geographical differences might be biased towards studies included through the search.

The absence of data on other anthropometric measures, such as Mid Upper Arm Circumference (MUAC), is also a potential limitation. In considering the implications of the differences highlighted here, in addition to biological and social explanations, it is necessary to consider how we measure and define undernutrition and whether sex differences are an artefact of the indices in use. The WHO growth standards describe the physiological growth within optimal environmental conditions and are separated by sex. This reference data from healthy well-nourished populations resolves sex differences to zero by expressing data as z-scores calculated using the appropriate male and female subset of the reference population. However, it is unclear if we would expect sex differences in undernutrition expressed in this way to be zero, when the distribution of weight and height in both sexes has been shifted away from the healthy reference range. Likewise, it is unclear if the loss of the same amount of body weight in a girl or boy would have the same physiological effect. If boys cope worse than girls when exposed to food shortages or disease and infection, this potentially highlights increased vulnerability over and above what is already accounted for by the standards.

In comparison, MUAC cut offs are unadjusted and do not differentiate by sex, or age (between 6 months and 5 years). This absence of adjustment may lead to a preferential inclusion of girls in programmes compared to what would be obtained if sex specific standards were used as girls tend to have lower MUACs than boys. Though it has been shown to be a good predictor of mortality, sex differences in using MUAC to define undernutrition have not been widely studied.

Finally, the number of studies identified in the overall search that qualified for the meta-analysis was low. This was mainly due to a lack of presentation of disaggregated data. A recent Lancet series on gender equality, norms and health, highlighted the need for accurate disaggregated data. [35]

### Implications for future research

This study is a step towards better understanding of sex differences in undernutrition and highlights the need to consider potential implications for policy and practice. Future research should aim to unpack the complexities related to age, biological and social risks (including gender norms) and context, and further our understanding of sex differences in the context of undernutrition and sub-regional variations in order to determine the implications of these differences for programme staff and policy makers.

Future research will focus on a more detailed analysis of factors affecting outcomes for boys and girls such as epidemiological, demographic and social differences, explore the consequences of sex, age, and behavioural differences in nutritional outcomes and mortality. The impact of using differing anthropometric measurement and indices should also be explored to better understand how differing methods detect the most vulnerable children and explore how substantial sex differences are.

## Conclusion

This review demonstrates that undernutrition defined by anthropometric case definitions is usually higher among boys than girls. Whilst further research is needed to understand the policy and programming implications of these differences, lessons can already be drawn from this research. We call on nutrition actors to improve data collection in programmes, surveys and research through the full disaggregation and analysis of sex and age in order to identify which children are most vulnerable in specific contexts, and to allow comparison of programme data with population level burdens. It is important to understand that the message of this study is not that boys should be prioritised over girls, rather it seeks to support all at-risk children, through improved understanding of sex differences in undernutrition. Ultimately, we believe all children under five years and their caregivers should be seen as a high priority group for targeted nutrition interventions and resources and interventions should be targeted according to need.

## Data Availability

All data reported in this paper are publicly available

## Acknowledgements

We would like to acknowledge and thank Zoe Thomas at the London School of Hygiene and Tropical Medicine Library for her review of the search terms used in this study.

